# IMAGINATOR 2.0: Co-design and early evaluation of a novel blended digital intervention targeting self-harm in young people

**DOI:** 10.1101/2025.03.27.25324316

**Authors:** Athina Servi, Emily Gardner-Bougaard, Saida Mohamed, Aaron McDermott, Nusaybah Chowdury, Rachel Rodrigues, Ben Aveyard, Nejra Van Zalk, Adam Hampshire, Lindsay H Dewa, Martina Di Simplicio

## Abstract

**Background:** Self-harm (SH) affects around 20% of all young people in the UK. Treatment options for self-harm remain limited and those available are either non-specific or long and costly and may not suit all young people. There is an urgent need to develop new scalable interventions to address this gap. IMAGINATOR is a novel imagery-based intervention targeting self-harm initially developed for 16- to 25-year-olds. It is a blended digital intervention delivering Functional Imagery Training (FIT) via therapy sessions and a smartphone app. In this study, we piloted a new version of IMAGINATOR extended to adolescents from age 12 after co-producing a new app with a diverse group of young people experts-by-experience. Here we report on feasibility of delivering IMAGINATOR 2.0 in secondary mental health services and clinicians’ feedback on the intervention.

**Methods:** Participants were recruited from West London NHS Trust Tier 2 CAMHS and adult Mental health Integrated Network Teams (MINT) teams. They received three face-to-face FIT sessions in which the app was introduced followed by five brief phone support sessions. Outcome assessments were conducted after completing therapy, approximately 3 months post-baseline. Two focus groups were conducted to gather the therapists’ perspectives on the IMAGINATOR 2.0 intervention. For quantitative data, descriptives are reported. Qualitative data were analysed using a co-produced thematic analysis method with lived experience co-researchers.

**Results:** Eighty-three participants were referred and 29 (28 female, 1 transgender, mean age = 18.9) were eligible and completed screening. Out of 27 participants who started, 59% completed therapy per protocol while only 15 completed the quantitative outcome assessment. There was an overall reduction in number of SH episodes over 3-months from pre-to post-intervention (baseline: median = 6.5, IQR = 35; post-intervention: median = 0, IQR = 7; median diff = -6.5, r = 0.69). Five themes were identified through analysis of therapists’ feedback, including therapy impact, mental imagery efficacy and limitations and need for better integration of the IMAGINATOR 2.0 app with therapy sessions. The app was overall well received, and therapists highlighted the need for an intervention like IMAGINATOR in their services.

**Conclusion:** We show that IMAGINATOR can be extended to adolescents, is acceptable and has potential as a brief intervention reducing self-harm in young people under mental health services. A future RCT is needed to robustly test the intervention efficacy, after considering issues around high attrition in outcome measures.

## INTRODUCTION

Self-harm (SH) is defined as the “intentional act of self-poisoning or self-injury, irrespective of the motivation” and is an expression of emotional distress (NICE, 2013) that typically begins in early adolescence, peaking in frequency around age 16 years (Gillies et al., 2018). SH prevalence has increased over the past 20 years across age groups in the United Kingdom (UK) (Gillies et al., 2018), with approximately 20% of individuals aged 16-25 report having self-harmed at some point in their life (Pisinger et al., 2019).

Despite the impact on young people’s lives, SH behaviour has been neglected by policy and research (Moran et al., 2024). Available targeted interventions are inadequate to the high level of need and better-quality evidence-base has been repeatedly called for (Ougrin et al., 2015; Witt et al., 2024). Interventions with the strongest evidence support (such as Cognitive Behavioural Therapy (CBT) (Witt et al., 2021) or Dialectic Behavioural Therapy (DBT) (Mehlum et al., 2014)) are either limited to adults or long, costly and requiring a high commitment. Therefore, we lack a stepped model of care starting with a brief early intervention for SH in young people prior to long, complex treatments. Digital solutions could be crucial to reach large numbers of young people and provide early support (Glenn et al., 2019) and are generally favoured by this population (Čuš et al., 2021). However, only a handful of app/web-based interventions for SH focus on and were co-designed with young people, and have been adequately tested (Bjureberg et al., 2023; Moran et al., 2024; Stallard et al., 2024).

### The role of Mental Imagery in Self-harming Behaviour

SH cognitions are often experienced in the form of mental imagery: vivid, realistic representations that can result in re- or pre-experiencing the same emotional and physiological reactions as the actual act (Ji et al., 2021). SH mental imagery has been identified in clinical (84% prevalence) (Lawrence et al., 2023) and student populations (73% prevalence) (Hasking et al., 2018) and has been shown to increase the urge and likelihood of future self-harm (Cloos et al., 2020; Ji et al., 2024; Wetherall et al., 2018). On the other hand, mental imagery of adaptive actions can promote engagement in the behaviour (Renner et al., 2019) and mental imagery-based interventions could help individuals in identifying and putting in place alternative strategies to replace SH.

Recent studies show that mental imagery interventions such as Functional Imagery Training (FIT) can be more effective than Motivational Interviewing at promoting change of problematic behaviours (Solbrig et al., 2019). FIT (Kavanagh et al., 2014) involves training and rehearsal of motivational imagery to enhance the desire of reaching adaptive goals (May et al., 2015). For example, it has been shown to promote weight loss (Solbrig et al., 2019) and reduce alcohol consumption (Shuai et al., 2021). IMAGINATOR is an imagery-based intervention delivering FIT to target SH via a combination of face-to-face therapy sessions and a smartphone app. In a proof-of-concept study, we showed that IMAGINATOR reduced SH after 3-months, which was maintained at 6-months in young people 16-25 years old (Di Simplicio et al., 2020).

Building on these results, we co-designed a new version of the app for adolescents and young adults aged 12-25 years old (IMAGINATOR 2.0) and aimed to assess its feasibility, acceptability and safety delivered in children/adolescent and adult secondary mental health services. Secondary objectives were to examine change in key outcomes, such as number of SH episodes, urges and mental imagery, other mental health symptomatology, and explore clinicians’ perspectives in delivering IMAGINATOR 2.0. Young people participants’ views are reported in a separate paper.

## METHODS

### Design

This was a single-arm study delivering FIT as a low intensity intervention alongside standard care in a sequential sample of young people (YP) under the care of West London NHS Trust (WLT).

### Co-production

A Young People Advisory Group (YPAG) of 4 young people with lived experience of SH was involved in funding application, design, recruitment, data collection, data analysis, interpretation of results, and dissemination. The YPAG shared decision-making authority with the research team throughout the project and all members co-led the the app co-design with the rest of the research team. One member was not able to stay involved in the later stages of the research due to ill health; one member (SM) conducted 3 feedback interviews with participants from the study; and three (SM, AM, NC) took part in a 3-hour thematic analysis training and in 6 thematic analysis meetings between January and April 2024.

### Co-design of the IMAGINATOR 2.0 App

We co-designed a new app with a diverse group of 14 YP with lived experience of SH recruited via social media across the UK, but with greater representation from Northwest London areas: age: 14-25, gender: 43% female/14% male/43% other, sexual orientation 22% heterosexual/78% other, ethnicity: 43% White/36% Asian/21% Black, 93% low to middle income. We conducted 4 structured half day co-design workshops following the Design Council’s Double Diamond Process, followed by one-to-one iterative usability testing of the app prototype, as described in a <u>case study</u> by the UX designers. Key functionalities selected for the final app included a degree of personalization, segmentation, inclusivity, and modularity (e.g., mascot to facilitate engagement, goals tracker, mood tracker, journal) (Figure 2-4). The app was coded by H2CD Ltd. Reflections from co-designers on the process highlighted feeling valued and listened to, appreciating the variety of activities, enjoying the opportunity to learn from diverse perspectives and experiences, and gaining insights into research and app design.

**Figure 2:**
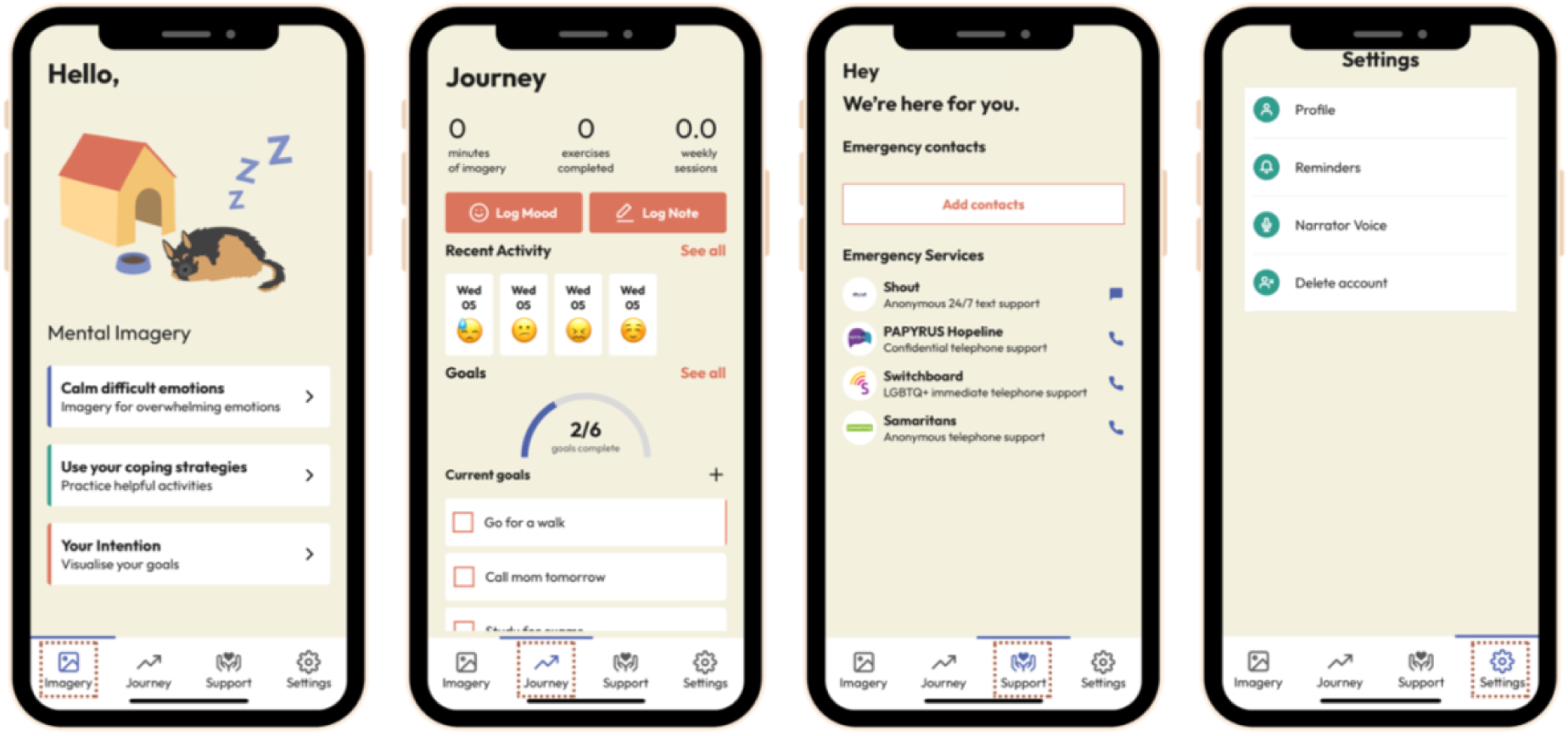
Main pages and appearance of the IMAGINATOR 2.0 app. The homepage of the app contains all imagery audios for YP to practice. The app also includes a tab where the YP can track their journey (mood, notes, goals) and available support where they can also add personal phones from people in their contacts. YP have the option to enable reminders for practicing mental imagery and logging mood choosing the time and days they want.

**Figure 3:**
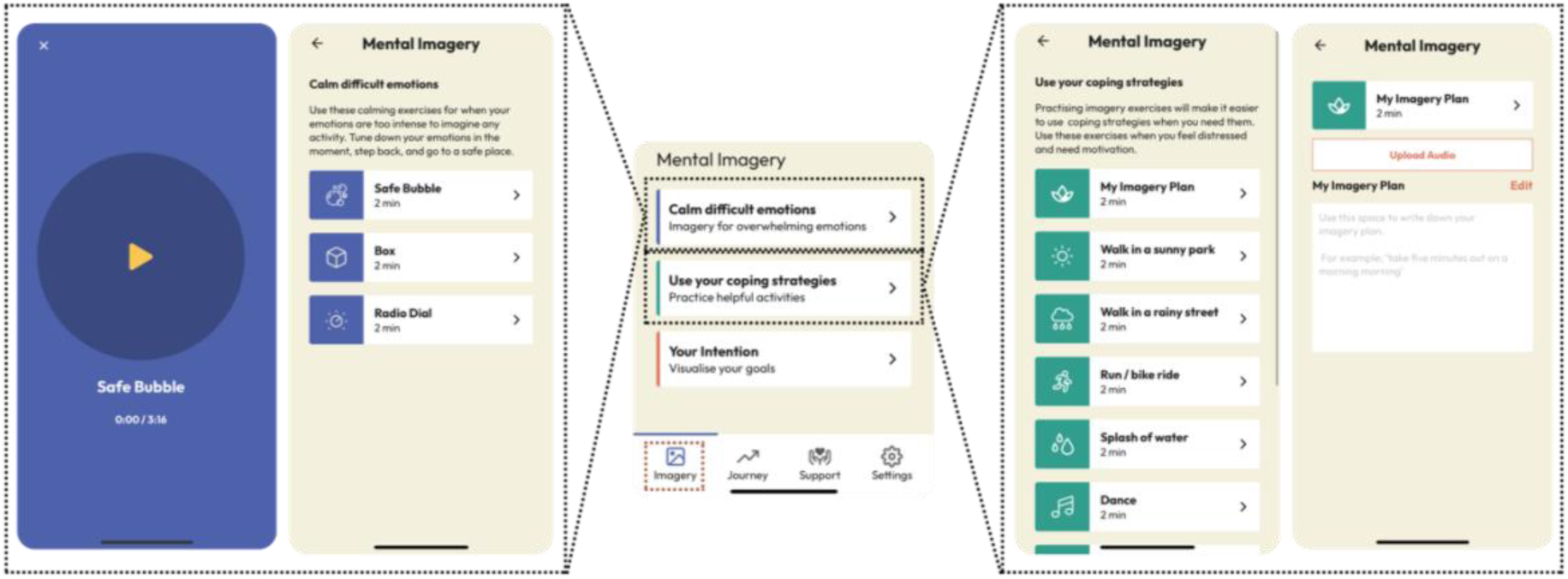
Mental Imagery in the IMAGINATOR 2.0 app. YP have a variety of option to practice mental imagery in the app that are accessible anywhere. They also have the option to record and upload their own imagery plan.

**Figure 4:**
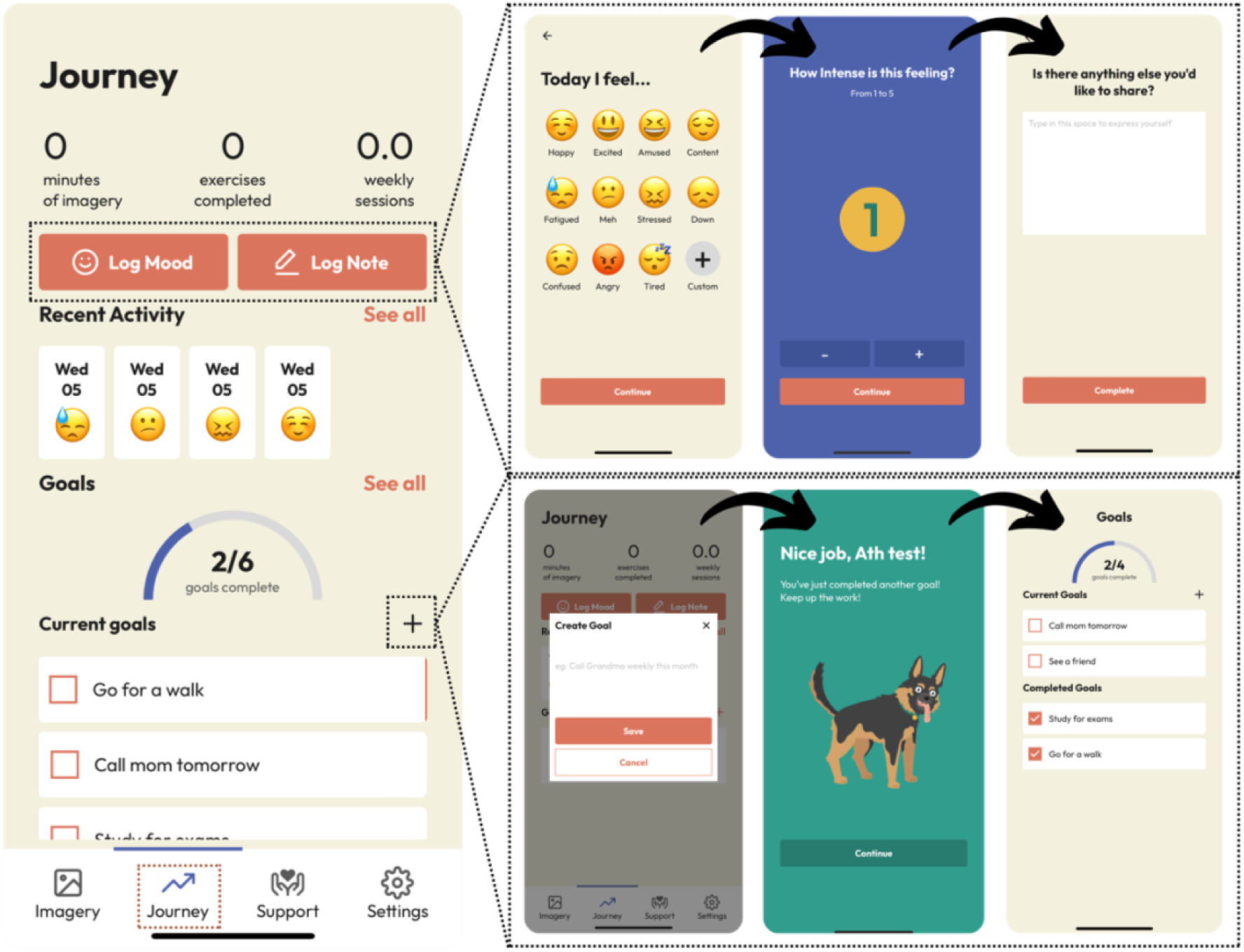
Logging mood and tracking goals in the IMAGINATOR 2.0 app. YP can track their mood every day, multiple times a day and rate it from 1 to 5 while also adding a note either for their mood rating or general. YP can also add their goals and check them off as they complete them.

### Participants and recruitment

Participants were recruited from CAMHS Tier 2 services (delivering low intensity psychological interventions) and MINT (adult community) teams in WLT. Participants were eligible if they had experienced at least 2 SH episodes lifetime with at least 1 in the past month, or at least 5 SH episodes in the past year and currently reporting self-harm urges; had a smartphone; were fluent in English; willing for parent/guardian to provide consent for study participation if 12-15-years-old. Participants were excluded if they had a severe learning disability or pervasive developmental disorder; current acute psychotic episode; current substance dependence; imminent risk of suicide or harm to others (based on clinicians ‘risk assessment); taking part in concurrent treatment studies for self-harm or in concurrent psychological therapy. Eligible participants were assigned a therapist from the referring team (Children and Wellbeing Practitioners; CWP, Clinical Assistant Psychology Therapists; CAPT, or Graduate Mental Health Workers; GMHW).

### Procedures

The study was approved by the West of Scotland Research Ethics Committee 5 (REC ref: 22/WS/0087). Recruitment took place over 9 months from November 2022 to September 2023. Potential participants completed a baseline screening either in person or via MS Teams video call, including assent/consent taking, sociodemographic characteristics (age, gender, ethnicity, sexual orientation, education, employment, and socio-economic status) and the measures listed below using Qualtrics online surveys.

### Intervention

IMAGINATOR consisted in 3 x 1-hour FIT sessions in person or via MS Teams once a week followed by 5 x 15-30-minute phone-call support sessions fortnightly. FIT comprised of the following key elements: Session 1 - formulation of personal drivers of SH behaviour and motivational interviewing combined with mental imagery techniques; Session 2 - mental imagery of a past achievement, goals setting (e.g., a desired behaviour incompatible with or alternative to SH) and functional imagery plan to achieve them; Session 3 - practice and refinement of functional imagery, problem-solving of challenges. The IMAGINATOR 2.0 app was introduced in Session 2 to support practicing helpful mental imagery at home. Follow-up phone-call sessions focused on refining functional imagery practice, problem-solving, motivational encouragement, and personalization of the app use (Figure 5). The therapist also conducted a risk assessment at the beginning of each session.

**Figure 5:**
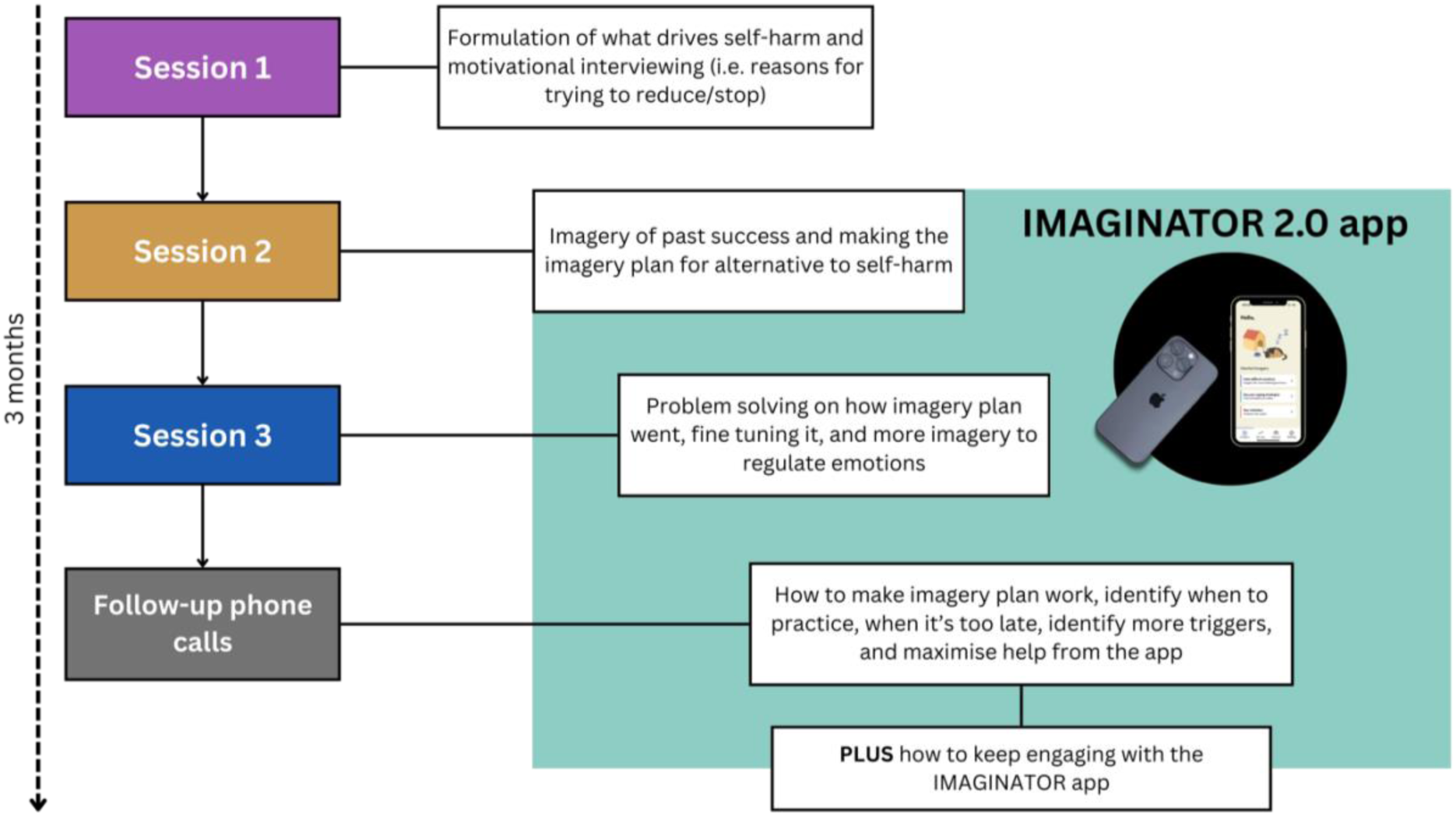
Overview of IMAGINATOR 2.0 intervention FIT sessions.

Therapists were trained in the delivery of IMAGINATOR 2.0 over two workshops (1 and ½ day in total), provided with a manual and supported via weekly group supervision with the study PI (MDS) and the Research Assistant (AS).

*Outcome Assessments*. Participants were invited to the outcome assessment after the final therapy session (approximately 3 months from baseline) via MS Teams video call and followed the same procedures and measures as the baseline assessment; then were invited to separate feedback interviews about their experience using IMAGINATOR 2.0 (reported in Dewa et al., in preparation).

### Focus groups

Therapists were invited to discuss their experience in delivering IMAGINATOR 2.0 across two focus groups that took place in person on 08/12/2023 and on 13/12/2023 at Imperial College London for approximately 2 hours, facilitated by the PI (MDS) and another researcher (AS and EG). A semi-structured interview guide with open-ended questions co-produced with the YPAG was used for the discussions, covering topics such as barriers to adopting the intervention, perceived benefits, and suggestions for improvement. The sessions were audio-recorded with therapists’ consent, transcribed verbatim, and anonymised. Field notes were also taken by the assisting researchers (AS and EG) to capture non-verbal cues and group dynamics.

### Measures

*Primary Outcomes* were, for feasibility, attrition (percentage of enrolled eligible participants completing outcome assessment) and treatment adherence (percentage of participants completing intervention per protocol); for acceptability, the *Client Satisfaction Questionnaire (CSQ)* (Attkisson & Zwick, 1982) and the *User Experience Questionnaire (UEQ)* (Laugwitz et al., 2008). Adverse events were recorded at each assessment and therapy session.

*Secondary Outcomes* included change in SH frequency and severity over the past 3 months, and in other SH-related and mental health symptoms assessed on measures below.

### Self-harm measures

- Timeline follow-back interview (TLFB; Sobell & Sobell, 2000): method conducted by the researcher who reconstructs together with the participant the number of self-harm behaviours using calendar cues, as used in previous proof-of-concept study (Di Simplicio et al., 2020).
- Self-harm severity over the past 3 months: a combined measure of a Visual Analogue Scale, severity criteria used in the NSSI disorder in the Diagnostic and Statistical Manual of Mental Disorder Fifth Edition (DSM-V, 39) and the number of different self-harm methods used (also termed self-harm versatility). Elicited via these open questions: ‘‘In the last three months, how severe was the worst injury that you inflicted to yourself? Can you list all the ways that you have used to harm yourself in the last three months?’’ This method was advised by the YPAG to avoid scales listing numerous self-harm methods which could be triggering/enabling.
- Self-Harm Imagery Interview (SHII) adapted from (Hales et al., 2011): a measure mental images of self-harm including PANAS ratings of affect following the image and other characteristics (e.g. vividness, controllability) on a 9-point Likert scale (Di Simplicio et al., 2020).
- Columbia-Suicide Severity Rating Scale (C-SSRS) (Posner et al., 2011): for suicide risk assessment and measurement of suicidal ideation and behaviour.
- State Motivation for Reducing Self-harm (SM-SH) scale (Parham et al., 2017): measures participants’ motivation to control their self-harm using 12 items rating the strength of motivational cognitions in the present moment on a 10-point Likert scale, from 0 (never) to 10 (constantly).
- Craving Experience Questionnaire for Self-Harm (CEQ-SH), adapted from CEQ (May et al., 2015): assesses the urge to self-harm as frequency, intensity, salience or dismissability of intrusive thoughts surrounding self-harm on a 10-point Likert type scale ranging from not at all (0) to constantly (10).
- *Mental health measures* were the Revised Children’s Anxiety and Depression Scale (RCADS) for 12 – 17-year-olds (Chorpita et al., 2015),
- Depression, Anxiety and Stress Scale (DASS-21) (Antony et al., 1998), for 18 – 25-year-olds,
- Warwick-Edinburgh Mental Well-being Scale (WEMWBS) (Tennant et al., 2007),
- Difficulties in Emotion Regulation Scale-Short Form (DERS-SF) (Kaufman et al., 2016),
- 11-item behaviour supplement to the Borderline Symptom List (Bohus et al., 2001),
- Alcohol Use Disorders Identification Test (AUDIT) (Saunders et al., 1993) and
- Cannabis Use Disorder Identification Test Revised (CUDIT-R) (Adamson et al., 2010)

### Analysis

#### Quantitative data

Given the feasibility nature of the study all quantitative data are reported as descriptive statistics and effect sizes with medians and interquartile ranges (IQR) as data was not normally distributed and effect sizes calculated as *r* = *z*/*sqrt*(*N*). Attrition, adherence to therapy per protocol, and number of therapy sessions completed are reported as percentages. To explore the relationship between adherence to therapy and various demographic variables, we conducted a Mann Whitney U test comparing the average number of therapy sessions completed between age, ethnicity and sexual orientation groups.

Due to the small sample size, missing data were tolerated, and we assumed that the data were missing completely at random.

#### Qualitative data analysis

Focus groups transcripts were reviewed and coded independently by two researchers (AS and EG). Data were analysed using a co-produced thematic analysis approach based on Braun & Clarke (2006) and Dewa et al. (2021). A Trello board was used to collate the initial codes separately added by both researchers so that all team members could review these. These were all incorporated into a coding framework. The research team (MDS, LD, AS, EG) and the co-researchers (SM, AM, NC) met twice online to review, refine, and finalise codes and one final time to co-produce the thematic map on Miro.

## RESULTS

### Sample

Sample demographics are reported in Table 1. Clinical characteristics are reported in Table 2 and 3. Among all participants, 80% reported experiencing suicidal ideation in the past month (assessed on the C-SSRS) with 60% reporting active suicidal thoughts and 13% a specific plan and intent to act on the plan; 16.7% of participants reported a lifetime history of suicidal attempts.

**Table 1:** Demographic characteristics of eligible participants at baseline assessment (*N* = 29).

| Variable |  | Mean (SD) |  |
| --- | --- | --- | --- |
| Age |  | 18.86 | (3.74) |
|  |  | Frequency (%) |  |
| Gender | Female | 28 | (96.55) |
|  | Transgender | 1 | (3.45) |
| Sexual Orientation | Heterosexual | 12 | (41.38) |
|  | Other | 12 | (41.38) |
|  | Homosexual | 3 | (10.34) |
|  | Not sure | 2 | (6.90) |
| Ethnicity | White | 17 | (58.62) |
|  | Black, African, Caribbean or Black British | 4 | (13.79) |
|  | Mixed or Multiple ethnic groups | 3 | (10.34) |
|  | Other ethnic group | 3 | (10.34) |
|  | Asian of Asian British | 2 | (6.90) |
| Household income | <50,000 | 18 | (62.07) |
|  | 50,000 – 100,000 | 9 | (31.03) |
|  | 100,000 – 150,000 | 1 | (3.45) |
|  | 150,000 – 200,000 | 1 | (3.45) |
| Highest level of education completed | Secondary education | 13 | (44.83) |
|  | Primary education | 8 | (27.59) |
|  | Higher education | 7 | (24.14) |
|  | Postgraduate education | 1 | (3.45) |
| **Currently in education training** | No | 15 | (51.72) |
|  | Yes | 14 | (48.28) |
| **Employment status** | No | 15 | (51.72) |
|  | Yes | 14 | (48.28) |

\*Standard deviation (SD)

**Table 2:** Change in self-harm related measures over 3-months after IMAGINATOR 2.0.

|  | Baseline (T0) |  | Outcome Assessment (T1) |  | Median Difference | Effect size (r) |
| --- | --- | --- | --- | --- | --- | --- |
|  | Median | IQR | Median | IQR |  |  |
| CEQ | 59 | 26.5 | 40 | 31 | -19 | 0.65 |
| SM-SH | 5.33 | 1.73 | 6.33 | 2.12 | 1 | 0.59 |
| SHII (PANAS+) | 14 | 9 | 16 | 5 | 2 | 0.02 |
| SHII (PANAS-) | 33 | 11.5 | 36 | 10.5 | 3 | 0.04 |

\*TLFB; Timeline Follow back Technique, CEQ; Craving Experience Questionnaire, SM-SH; State Motivation for Reducing Self-Harm, SHII; Self-Harm Imagery Interview, PANAS;
Positive and Negative Affect Schedule.

**Table 3:**
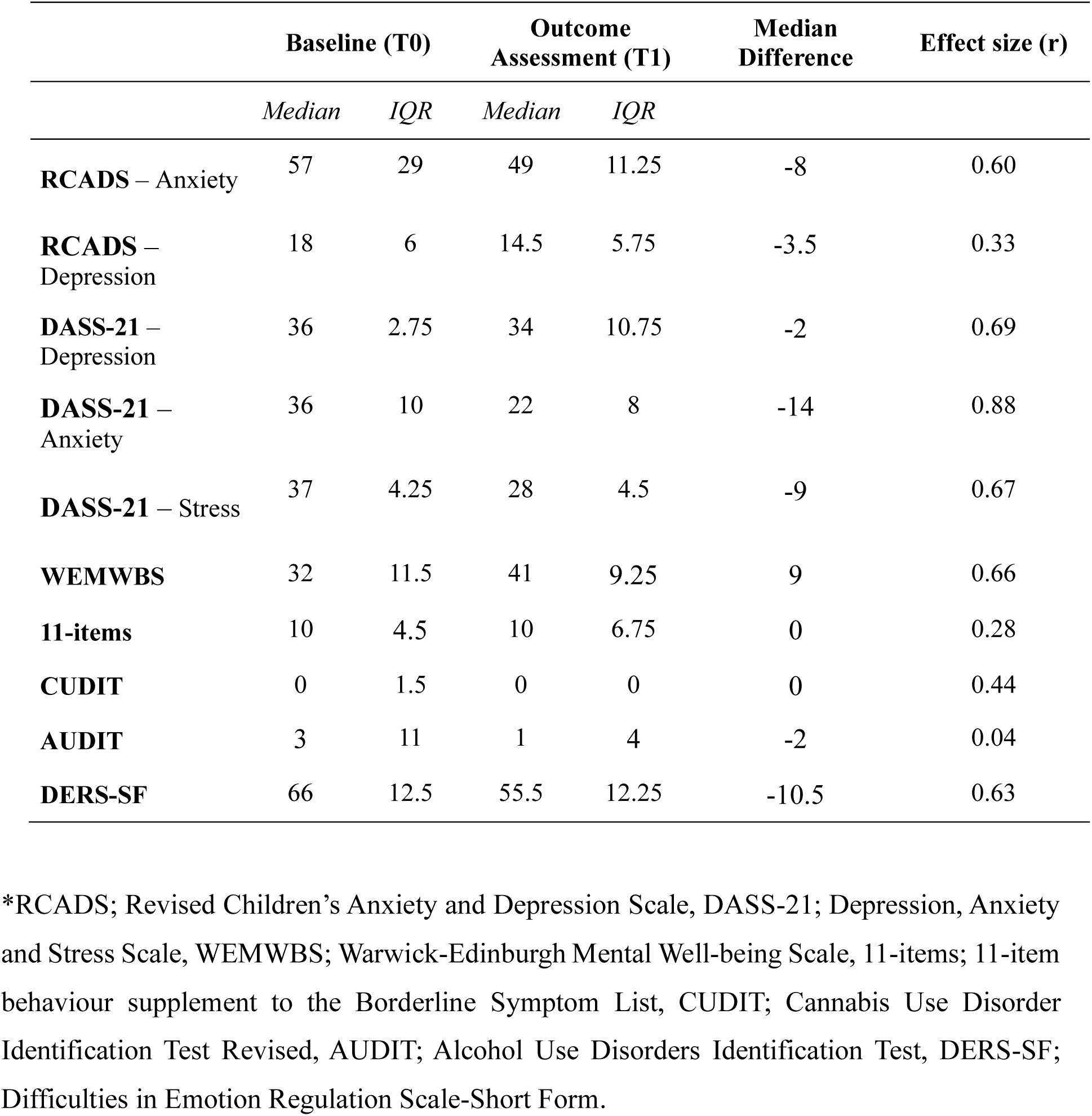
Change in mental health and wellbeing measures over 3-months after IMAGINATOR 2.0.

|  | Baseline (T0) |  | Outcome Assessment (T1) |  | Median Difference | Effect size (r) |
| --- | --- | --- | --- | --- | --- | --- |
|  | Median | IQR | Median | IQR |  |  |
| **RCADS – Anxiety** | 57 | 29 | 49 | 11.25 | -8 | 0.60 |
| **RCADS – Depression** | 18 | 6 | 14.5 | 5.75 | -3.5 | 0.33 |
| **DASS-21 – Depression** | 36 | 2.75 | 34 | 10.75 | -2 | 0.69 |
| **DASS-21 – Anxiety** | 36 | 10 | 22 | 8 | -14 | 0.88 |
| **DASS-21 – Stress** | 37 | 4.25 | 28 | 4.5 | -9 | 0.67 |
| **WEMWBS** | 32 | 11.5 | 41 | 9.25 | 9 | 0.66 |
| **11-items** | 10 | 4.5 | 10 | 6.75 | 0 | 0.28 |
| **CUDIT** | 0 | 1.5 | 0 | 0 | 0 | 0.44 |
| **AUDIT** | 3 | 11 | 1 | 4 | -2 | 0.04 |
| **DERS-SF** | 66 | 12.5 | 55.5 | 12.25 | -10.5 | 0.63 |

\*RCADS; Revised Children’s Anxiety and Depression Scale, DASS-21; Depression, Anxiety and Stress Scale, WEMWBS; Warwick-Edinburgh Mental Well-being Scale, 11-items; 11-item behaviour supplement to the Borderline Symptom List, CUDIT; Cannabis Use Disorder Identification Test Revised, AUDIT; Alcohol Use Disorders Identification Test, DERS-SF; Difficulties in Emotion Regulation Scale-Short Form.

### Feasibility and acceptability

Eighty-three patients were referred to the study, of which 29 were both eligible and completed baseline screening (Figure 1). Out of the 29 participants who were enrolled in the study and completed the baseline assessment, 27 (93.10%) started therapy. Out of the 27 who started therapy, 16 (59.3%) adhered to therapy per-protocol (all face-to-face sessions and at least 2 phone calls), 19 (70%) completed all face-to-face therapy sessions and 9 (33.3%) completed all five-follow-up phone-calls. On average, the waiting time to start the intervention was 4.1 weeks (*range*: 0.7 – 9), while the average intervention duration was 21.3 weeks (*range*: 9.7 – 36). Out of the 27 participants who started therapy, only 15 (51.7%) completed the follow-up outcome assessment.

The average number of therapy sessions completed was 4.93 (*SD* = 2.80) and it differed by sexual orientation with LGBTQ+ participants completing a higher number of sessions compared to heterosexual participants (*Mann-Whitney U test* = 39.500, *p-value* = 0.012). No other demographic variable such as age or ethnicity differentiated therapy adherence.

Participants reported a median score of 25 (*IQR* = 5.5) on the CSQ indicating a very good satisfaction of the intervention received. Ratings on the UEQ referring to the app indicated good levels of perspicuity (*Median* = 1, *IQR* = 1.5) and dependency (*Median* = 1.25, *IQR* = 1.63), acceptable levels of attractiveness (*Median* = 1, *IQR* =1.58), but low levels of efficiency (*Median* = 1.25, *IQR* = 1.63), stimulation (*Median* = 0.75, *IQR* = 1.5) and novelty (*Median* = 0, *IQR* = 0.63).

### Secondary Outcomes

SH episodes reduced from a *median* = 6.5 (*IQR* = 35) at baseline to *median = 0* (*IQR* = 7) after IMAGINATOR 2.0 across all participants (*effect size (r)* = 0.69, medium), see *Figure 7* in participants split by age group.

**Figure 6:**
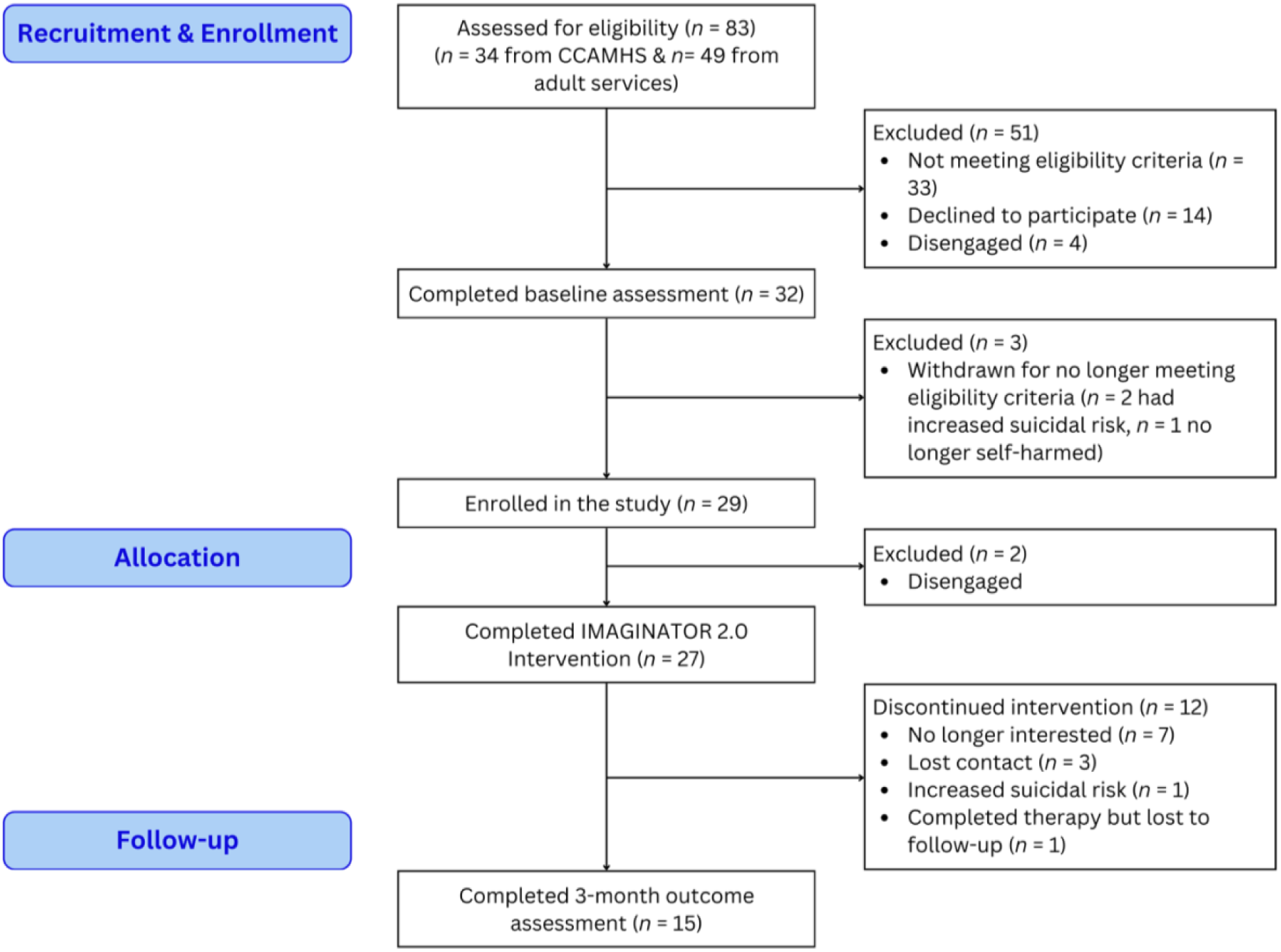
Flow diagram of participants in the IMAGINATOR 2.0 study.

**Figure 7:**
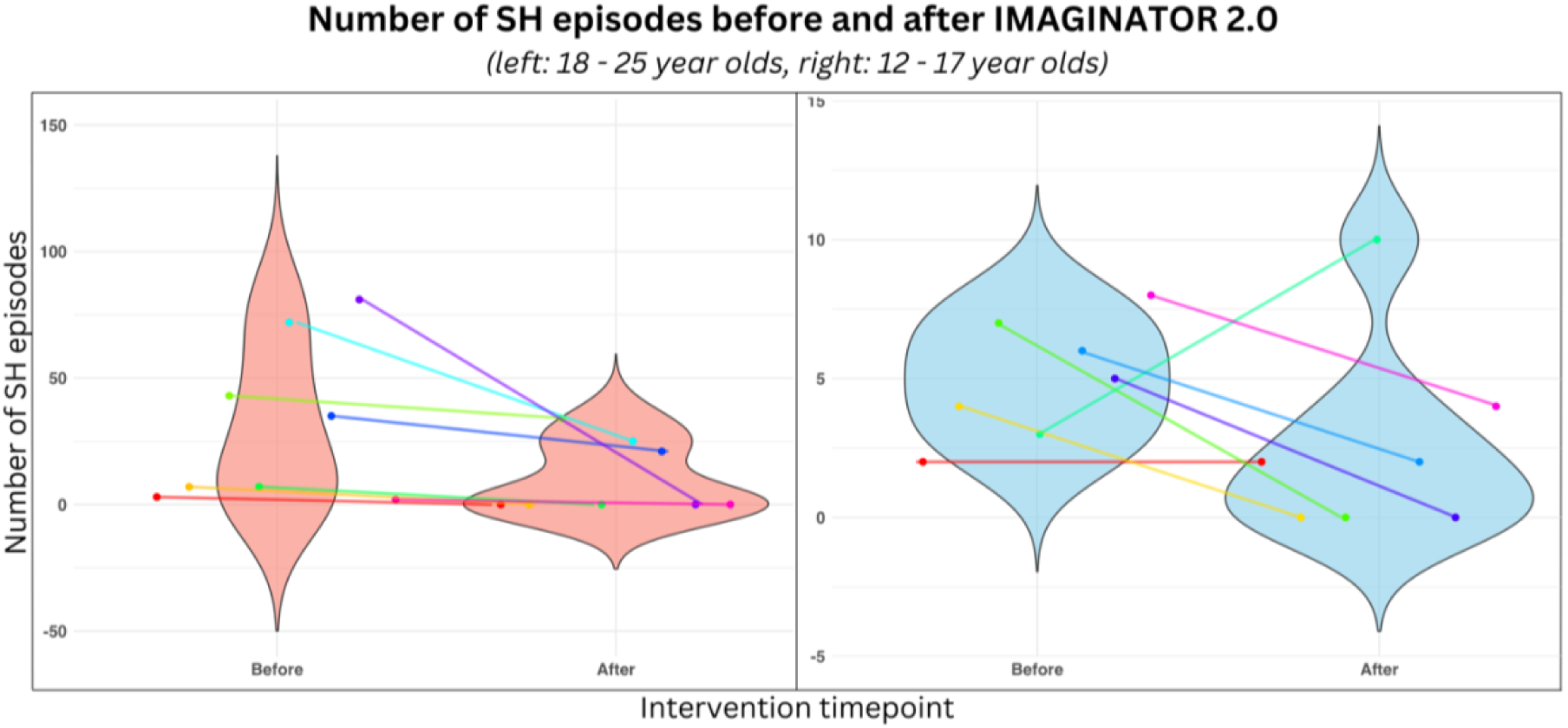
Number of SH episodes pre-and-post intervention in 15 participants who completed both baseline and outcome assessments, grouped by age into CAMHS (<18 years old) and MINT (≥18 years old) teams. Colour coded for each participant.

IMAGINATOR 2.0 effects on other clinical secondary outcomes are reported in Table 2 and 3. In terms of SH, IMAGINATOR 2.0 reduced craving for SH and increased participants’ motivation to reduce SH. IMAGINATOR 2.0 also reduced depression, anxiety, and stress in participants aged 18 or over, and improved all participants’ wellbeing and emotion regulation skills.

There were no adverse events reported during the study.

### Qualitative analysis from focus groups

All 8 therapists took part in feedback focus groups: five from adults and three from adolescent services. Qualitative thematic analysis identified the following six themes: IMAGINATOR 2.0 therapy impact, mental imagery efficacy and limitations, therapist requirements and support to deliver therapy, app experience and engagement, potential changes and app integration, and IMAGINATOR 2.0 expectations and need for improvement. Connections between themes and sub-themes are conceptualised in a co-produced thematic map (Figure 8).

**Figure 8:**
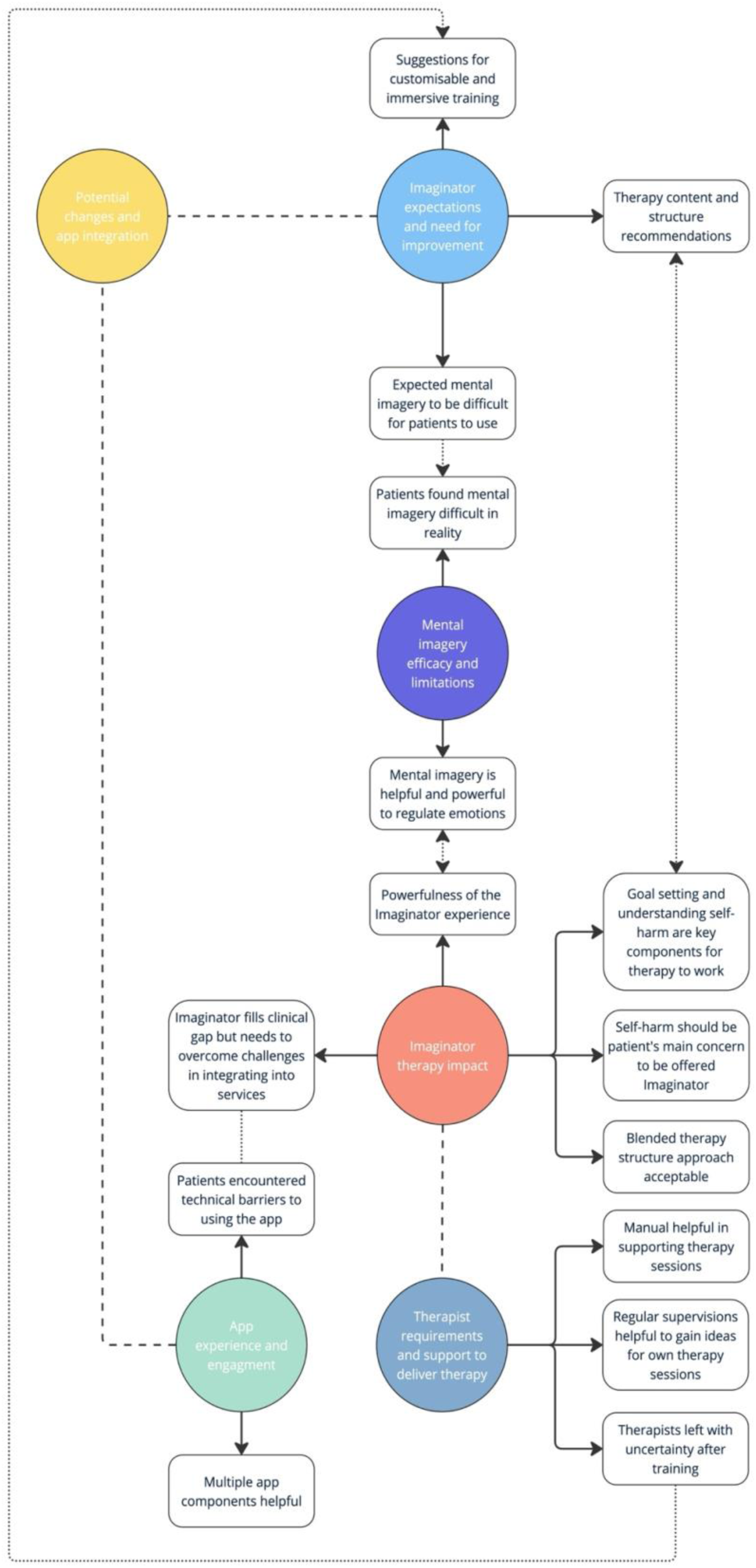
Co-produced thematic map of therapist focus group feedback.

#### IMAGINATOR 2.0 therapy impact

All therapists found that IMAGINATOR 2.0 was a powerful experience for patients and filled a critical gap in service provision offering a needed short-term intervention targeting SH for young people that could reduce waiting times for this group. However, preferably there should not be waiting for IMAGINATOR 2.0 itself and it should not preclude being on a waiting list for longer therapies if needed.

*“I think Imaginator was a really brilliant experience [..] Having kind of a short-term therapy to reduce self-harm has been really helpful for my participants and they said they found it really useful.”*

*(Therapist ID: 6)*

*“What’s good about IMAGINATOR is that it is focused on short term, which sometimes is what is needed in that moment just to manage that sort of self-harm escalation”*

*(Therapist ID: 1)*

*“At the moment in our team, the waiting list for DBT is over a year long, so people are left with actually no kind of options, no coping skills in between and they’re not offered anything.”*

*(Therapist ID: 6)*

*“For a lot of people, it’s their first opportunity to actually discuss self-harm in detail because in risk assessment when you ask that question it does not necessarily feel comfortable. But this intervention normalised it and gives a space to understand it.”*

*(Therapist ID: 2)*

Therapists agreed that patients referred to IMAGINATOR 2.0 should have the primary goal to change their SH behaviour, or they may disengage from the intervention. Some felt that age above 14 years old is preferable to make the concept of mental imagery easier to grasp; others that a longer history of self-harm could facilitate self-awareness and engagement. Understanding self-harm and goal setting were viewed as the key components of the therapy.

*“Self-harm can escalate even though people are involved with [mental health services]. So, participants need something to just sort of contain the self-harm and Imaginator can do that quite well.”*

*(Therapist ID: 1)*

*“I think understanding the self-harm and goal setting [are key components of the therapy] because that is what is going on and for them that was a big problem. Understanding what would be helpful for them.”*

*(Therapist ID: 3)*

*“I think probably older young people would benefit more. […] So, I think maybe for young people who have a longer history of self-harm and there’s sort of a clearer kind of relationship to their self-harm and they have already tried to do something about it.”*

*(Therapist ID: 8)*

Most therapists found the blended therapy approach (in-person, app and calls) a good structure catering for patients’ different needs. However, during phone calls patients were often distracted and these sessions were not as efficient as face-to-face.

*“Overall, I sort of like the structure. I think it’s sort of slowing down and basically having a repeated structure towards the end makes sort of the ending easier”*

*(Therapist ID: 2)*

*“It’s just chaotic because you can hear in the background, they are playing video games or they’re doing something else […] So I think that’s a normal experience of phone calls with young people. And I think that’s probably not as helpful for Imaginator”*

*(Therapist ID: 1)*

#### Mental imagery efficacy and limitations

All therapists highlighted the powerfulness of mental imagery as an emotion regulation technique; even if this was not always sufficient for all young people and for some it was difficult to practice. Another key strength of mental imagery was the freedom to practice it anywhere at any time. Therapists also valued the collaborative process of trial-and-error with the patient to identify the most helpful adaptive image and many reported that they were likely to adopt mental imagery techniques in their future routine sessions.

*“One young person’s goals had more to do with social anxiety. They seemed to find it very useful to picture themselves in situations that they expected to find challenging and to actually vividly imagine themselves coping and having worked out what they would do in these situations. They found that helpful. It was a very collaborative process*.

*(Therapist ID: 8)*

*“One of my participant’s goals was to reduce her negative self-talk because she puts herself down a lot. She found it really helpful and powerful to imagine some of the things that she should be saying to her younger self because she was thinking that that was an innocent person. This helped her to blow off some of the guilt and shame that she had about herself in the present and by associating herself with her younger self through imagery really helped her*.

*(Therapist ID: 7)*

*“I think for me often with coping tools outside imagery, the young people feel they have to be in a certain place, or they have to sort of have a certain object or thing, whereas mental imagery is so accessible, and can literally just be used anywhere.”*

*(Therapist ID: 4)*

*“One of my participants sometimes found it hard to kind of stay on the image that she wanted. And her mind started to wander. And it started to go to negative places.”*

*(Therapist ID: 7)*

#### IMAGINATOR 2.0 expectations and need for improvement

Most therapists expressed initial hesitation around mental imagery as a tool to reduce SH. There was also some scepticism around how to explain mental imagery and how it would work, and concern about the young age of patients being a barrier to understanding and using mental imagery techniques. For most, these negative expectations had dropped by the end of intervention.

*“I thought that imagery would be a difficult concept to grasp.”*

*(Therapist ID: 1)*

*“At the beginning, I was slightly sceptical, and I didn’t understand everything about the research. I was just wondering how helpful it could be and thinking in terms of imagery and how it would work. But then, after working with my first participant and recognising how she enjoyed it and how it was really working for her, I kind of changed the opinion I had initially.”*

*(Therapist ID: 7)*

*“So, the idea of using imagery seemed quite a compelling approach but I did find it quite challenging to deliver. Which I think was connected with the age group that I was working with.”*

*(Therapist ID: 8)*

Some therapists highlighted the need to add more focus directly on regulating emotions as well as a way to deal with SH urges. Another suggestion was to integrate IMAGINATOR 2.0 as part of routine safety planning and train more people from the team to deliver it as a SH relapse prevention measure.

*“Using mental imagery as more than just an ad hoc tool for managing self-harm during my therapy sessions, has actually been quite effective with young people [outside Imaginator]. When doing our routine safety plan, we discuss how they can use imagery. This could be part of our toolkit that we use in safety planning maybe.”*

*(Therapist ID: 1)*

*“I think there is definitely some usefulness [for Imaginator] in our team. We’ll have to think a bit about how it fits coherently and who would be delivering it and how they would be supported in doing that. I think there’s an interesting potential. […] I think there could be something about using imagery and incorporate all the members of the team to become more engaged and incorporating that in their work.”*

*(Therapist ID: 8)*

*“Maybe first they do the managing emotions group to learn how to deal with difficult emotions and then if self-harm is still an issue put them on Imaginator.”*

*(Therapist ID: 5)*

#### Therapists’ requirements and support to deliver therapy

Most therapists found the one-day workshop training informative but were left with uncertainty around what the actual therapy was going to look like. Some only understood aspects of the training when they finally conducted their first therapy session and mentioned the information could be overwhelming without sufficient previous therapy experience. Most therapists suggested splitting the training over two days, increasing role-plays, practice of mental imagery techniques and having video resources to refresh information.

*“I really enjoyed the training. The little consolidation training we did closer at the time, was really helpful because it was like more role play. So, seeing how you would actually use the app and having like visual example of that and how conversation could go even though we had the scripts, it was just easier to kind of see how it would be like in an actual session. That I found that really helpful.”*

*(Therapist ID: 7)*

*“In the training it was not so clear but then doing it for the first time helped in understanding.”*

*(Therapist ID: 3)*

All therapists found the manual practical and a useful tool to prepare for sessions.

*“It was nice to have that sort of side [the manual] and the sense of knowing what direction to take to session. Sometimes with my client I found that we followed the manual a little bit but it’s flexible in the sense that we don’t have to sort of stick to the manual exactly. So, it’s quite helpful to have it there.”*

*(Therapist ID: 5)*

All therapists found regular supervision sessions helpful, a safe space and a learning session via hearing other people’s experiences. When faced with a challenge, supervision allowed identifying alternative routes to how to approach a patient.

*“I wonder how much people are using imagery, but in CAMHS I suspect probably not very much. So, I think having this sort of space where we would think about it together and have that kind of supervision was quite helpful in terms of hearing others ‘experience.”*

*(Therapist ID: 1)*

#### App experience and engagement

Most therapists reported that the IMAGINATOR 2.0 app was user friendly and age appropriate. The audios were the app feature most accessed, while for some mood logging also appeared useful for young people. Therapists valued that the app had multiple components, but opinions varied around which was the key one: guided imagery audios, the support section or the goal section.

*“The participant I worked with she had quite a positive reaction to the app, and she used it between sessions as well. I think for her it was quite helpful because it reminded her to use imagery even outside of our sessions. So, I think she found the box audio quite helpful. Mood logging was helpful. She was able to see that she had more good days than she actually thought*

*(Therapist ID: 5)*

*“The audios where the most helpful for one of my participants. She didn’t use the app much, but she found audios helpful. So, the safe bubble and the other ones she did explore them in her free time a few times. I wouldn’t say it was regular, but she did listen to them.”*

*(Therapist ID: 6)*

Therapists noted that a few young people were excited to use an app. However, a major challenge were technical difficulties, which prevented young people from engaging further with the app outside therapy sessions, and a few patients abandoned the app because of these. Some patients found the voices/accents to be jarring and so did not use the audios. Few therapists believed that the app should have more personalisation to address patients’ needs and increase engagement.

*“Had some difficulties in encouraging young people to use it. The young people were quite picky about voices and sort of general bugs.”*

*(Therapist ID: 2)*

#### Potential changes and integration of app

Most therapists highlighted that the app needs to be more integrated into the therapy: this could be done by having one more dedicated face-to-face session, to guide patients through the app and reinforce its use.

*“We should sit down and see things together because sometimes they don’t do it on their own or they have difficulties with tracking mood etc. My young person was excited to use the app. Even though their initial reaction was positive, exploring it on their own did not work”*

*(Therapist ID: 3)*

*“I think it would be easier to start using the app or have things to do in the app from all the face-to-face sessions because it kind of consolidates it every week. So, if there’s some activity or task to do on the first session, in the second session, and then in the third session, and having them recording it, kind of sets them up for when they’re going off by themselves, and then we can consolidate it further when we’re doing the phone calls. I think it would make it a lot easier for them to not forget the app and actually kind of integrate it more into the module. Whereas right now it kind of feels like two separate things.”*

*(Therapist ID: 7)*

A few therapists thought that using an incentives system could encourage patients to use the app more. Making the app visible and accessible on the patient’s phone was important; improving the prompts/notifications system and for the therapist to record audios were other suggestions.

*“Thinking about just different avatars for themselves. So, people having their own kind of avatar that they could create, you know? As you would in a game with different codes and hair and all of that, that would be a fun option. Make it more personal to them…”*

*(Therapist ID: 6)*

## DISCUSSION

This study extends our previous proof-of-concept trial (Di Simplicio et al., 2020) of an imagery-based intervention in young people who SH showing that FIT is in principle well received, acceptable, safe and has potential at reducing SH behaviours in adolescents and young adults via a blended digital therapy approach. Here we also show reductions in associated psychopathology to various degrees. These results should be treated with caution given the small sample size and absence of a comparator intervention, which are key limitations in our study.

### Feasibility of IMAGINATOR 2.0

The high number of referrals (*N* = 83) from services highlights that IMAGINATOR 2.0 fills a gap in available interventions to support the high presentation of young people who SH (Trafford et al., 2023). More referrals from adult compared to adolescent teams may reflect the larger population covered (9 adult vs 2 community CAMHS teams) or that this need is higher for those aged > 18 years old (Diggins et al., 2017). Interestingly, adolescent clinicians requested that inclusion criteria be widened to young people with distressing SH urges even if they did not have a SH episode in the last month highlighting an overall lower threshold for intervention for this age group. In adult services instead, some young people would have been otherwise discharged from services as neither suitable to receive DBT, nor suitable to take part in emotion regulation skills groups due to recent SH or preference for individual therapy.

In line with our previous study (Di Simplicio et al., 2020) and other brief CBT interventions for SH (Slee et al., 2008; Yuan et al., 2019), adherence to face-to-face therapy was satisfactory (70%). Instead, attrition rate (48.3%) was higher than expected and compared to other fully digital interventions for SH which reported lower dropout rates (15% and 28%) (Bjureberg et al., 2023; Stallard et al., 2024). Most dropouts happened after the completion of the three face-to-face sessions, possibly suggesting that participants might have perceived phone call sessions as less beneficial or engaging compared to face-to-face interactions. Therapists emphasised how phone calls can be distracting for both participants and them, which is consistent with previous literature suggesting a lack of control of the environment during phone calls (Brenes et al., 2011). Future adaptations of our intervention could include an additional face-to-face session instead of calls or offer the flexibility of video calls to better maintain the therapeutic connection (Chen et al., 2022).

Most of our attrition reflected that participants disengaging from therapy after a few sessions were also lost to outcome assessment. This is frequently seen in psychological research (Alavi et al., 2013; Wei et al., 2013) hindering collection of feedback from participants who do not complete therapy and efforts to understand the reasons behind dropouts. In the final stage of our study, we noted that consistency in the research team conducting assessments and keeping in touch during the intervention with brief messages improved retention. Importantly, long waiting times between enrolment and therapy start might have led to change in needs (Punton et al., 2022; Quinlivan et al., 2023): e.g. some YP declined starting therapy as they had not self-harmed since the baseline assessment, while two adolescents experienced increased suicidal risk while waiting for therapy and were moved to a different team.

As emphasised by our therapists, SH has to be the patient’s main concern especially as ambivalence around reducing SH is often present (Gray et al., 2023) while not necessarily overt, and this ambivalence could persist during long waiting periods. The motivational interview in Session 1 may have self-selected those still in a contemplative phase of behaviour change explaining some early dropouts (Michaelsen & Esch, 2021). Additionally, by reducing the waiting time, therapy could better align with patient’s most pressing concerns. Cognitive markers of motivation to stop self-harming not relying on self-report may bypass the risk that young people start therapy because they feel that “they have to”, and lead to more personalised approaches (Rodrigues et al., 2023).

### Acceptability of IMAGINATOR 2.0

Both participants’ quantitative satisfaction ratings and therapists view indicate that our intervention was acceptable and well-received. The IMAGINATOR 2.0 app was judged reliable, intuitive to understand, and overall attractive both by young people and therapists. Young people’s low ratings of app’s stimulation are reflected in some therapists’ suggestions around adding gamification or further personalisation. Overall, this supports the idea that an app can enhance control and autonomy over one’s own care, which appears key in learning to manage SH behaviours (Hetrick et al., 2020). Integrating therapists’ suggestions in future app versions could improve this by sustaining engagement with the app.

Therapists strongly supported our blended digital model but recommend a more structured integration of the app in the therapy sessions. Importantly, this critique that app and face-to-face therapy remained too separate was also shared by young people’s interviews (Dewa et al., in preparation), possibly reflecting an ongoing challenge in maximising the potential of academic-led digital technology for therapeutic interventions with apps failing to show superior efficacy to standard care so far (Stallard et al., 2024).

### Mental Imagery

The perceived powerfulness of IMAGINATOR 2.0 appears directly aligned with the perceived power of mental imagery. Reductions in emotional dysregulation scores and craving to SH as well as higher motivation to stop confirm mental imagery as a cognitive process that can regulate emotions and support adaptive behaviours (Ji et al., 2019), notwithstanding the limitation of our single-arm design. Our findings are also consistent with growing evidence on the efficacy of targeting distressing involuntary imagery or harnessing the power of helpful imagery in anxiety and depression (Pile et al., 2021). Importantly, therapists’ view of the intervention and imagery in particular also match young people’s subjective experience describing that IMAGINATOR 2.0 produced an emotional and behavioural change, impacted on their ability to regulate emotions and behaviour, and improved motivation and goals-setting (Dewa et al., in preparation).

While the exact mechanism of FIT in the context of SH remains to be tested, it is possible that a personalised imagery plan simulating a way out of distress may work by diminishing the sense of entrapment, a recognised motivational factor in self-harm (O’Connor & Portzky, 2018). Alternatively, it could work by offsetting self-harm mental imagery (Ji et al., 2024). Another element that could explain this “powerfulness” is that the content of this imagery is highly personalised to each individual’s formulation and endorsed SH functions, from “reducing distress” to “feeling something” (Coppersmith et al., 2021; Nock et al., 2009), in line with recent guidance to approach self-harm in a more individualised way (Moran et al., 2024). Interestingly, therapists felt that key elements that made the intervention “work” were young people understanding their own SH formulation and improved goal-setting ability, but also described that the most used feature of the app were mental imagery audios.

Importantly, therapists initially anticipated that mental imagery would be difficult to implement especially in younger participants due to challenges with abstract concepts. Mental imagery techniques have found to be effective and doable in participants as young as 14-years-old (Schwarz et al., 2021; Schwarz & Stangier, 2023), but our therapists noted that those younger than 14-years-old often found it difficult. In fact, research suggests that vividness in mental imagery relies on both long-term and working memory, which may not fully developed before the age of 14 (Baddeley & Andrade, 2000; Kosslyn et al., 1990). This may explain why younger individuals struggled more with controlling and altering mental images consistent with their cognitive and emotional developmental stage (Burnett Heyes et al., 2013).

Therapists pointed out the challenges stressed the importance of regular rehearsal of the mental imagery plan via the app. A better integration between app and therapy may address this by specifically setting exercises that boost mental imagery’s motivational elements and via repeated pre-experiencing of positive emotions and desired outcomes. This would ensure participants enhance their ability to manage distress via imagery before reaching a crisis point level (Sosic-Vasic et al., 2024). Finally, therapists suggested that more direct targeting of emotion regulation *per se* would improve IMAGINATOR 2.0. This may reflect young people wanting to target underlying difficulties, e.g. emotional dysregulation, rather than caring about reducing self-harm (Knowles et al., 2022), and is supported by the recent evidence on the efficacy of ERITA, a web-based emotion regulation program, at reducing SH episodes (Bjureberg et al., 2023).

As mental imagery-based techniques appear to be under-used in particular by children and adolescent therapists, it will be important to develop more training including videos and interactive resources.

## CONCLUSION

In summary, IMAGINATOR 2.0 is the first blended digital intervention developed with and for young people who experience SH behaviour across adolescence and young adulthood, in line with NICE guidelines on the long-term management of self-harm (NICE, 2022). Our evidence suggests that it is safe, acceptable, and feasible to be delivered in mental health services to support young people with low levels of risk. Its brevity and low clinician input make it highly scalable across a variety of health services with the potential to be an early digital intervention to reduce SH in young people, filling the current gap in treatments for this population.

Future studies should test the efficacy of IMAGINATOR 2.0 in a randomised controlled trial, considering extending it to other services and to different community settings where young people initially seek help, such as schools, general practitioners and non-statutory services. Additionally, mechanistic knowledge could improve impact and guide personalisation, for example testing if the intervention works by reducing self-harm mental imagery and what app components directly influence treatment response. Finally, given its general focus on emotion regulation and behaviour change strategies IMAGINATOR 2.0 could be adapted as an early intervention targeting a variety of dysregulated behaviours.

## Data Availability

All data produced in the present study are available upon reasonable request to the authors

## ACKNOWLEDGMENTS

First, we thank all the young people who participated as co-designers of the IMAGINATOR 2.0 app for their invaluable critical enthusiasm and scepticism. Second, we thank UX designers Caylee Farndon-Taylor, Cui-Lyn Huang, Shalyn Wilkins, and Leroy Farndon-Taylor, and Pete Hellyer who coded the app for H2 Cognitive Designs Ltd for their dedicated work and ability to collaborate with many stakeholders. Finally, we thank all the therapists at West London NHS Trust for their interest and collaboration and the R&D team for their support with participant recruitment.

This project is funded by the NIHR i4i (NIHR203904). The views expressed are those of the author(s) and not necessarily those of the NIHR or the Department of Health and Social Care.

## ABBREVIATIONS

SH: Self-harm
CBT: Cognitive Behavioural Therapy
DBT: Dialectical Behavioural Therapy
FIT: Functional Imagery Training
YP: Young People
WLT: West London Trust
YPAG: Young People Advisory Group
CAMHS: Child and Adolescent Mental Health Services
MINT: Mental Health Integrated Network Teams
CWP: Children and Wellbeing Practitioners
CAPT: Clinical Assistant Psychology Therapists
GMHW: Graduate Mental Health Workers
CSQ: Client Satisfaction Questionnaire
UEQ: User Experience Questionnaire
TLFB: Timeline Follow-Back Technique
SHII: Self-Harm Imagery Interview
PANAS: Positive and Negative Affect Schedule
C-SSRS: Columbia-Suicide Severity Rating Scale
SM-SH: State Motivation for Reducing Self-Harm
CEQ-SH: Craving Experience Questionnaire for Self-Harm
RCADS: Revised Children’s Anxiety and Depression Scale
DASS-21: Depression, Anxiety and Stress Scale
WEMWBS: Warwick-Edinburgh Mental Well-being Scale
AUDIT: Alcohol Use Disorders Identification Test
CUDIT-R: Cannabis Use Disorder Identification Test Revised
IQR: Interquartile Range
DERS-SF: Difficulties in Emotion Regulation Scale-Short Form

## DATA AVAILABILITY

Data is available upon request.

